# SARS-CoV-2 infection- induced seroprevalence among children and associated risk factors during pre- and omicron-dominant wave, from January 2021 through November 2022, Thailand: Longitudinal study

**DOI:** 10.1101/2022.12.01.22283006

**Authors:** Nungruthai Suntronwong, Preeyaporn Vichaiwattana, Sirapa Klinfueng, Jiratchaya Puenpa, Sitthichai Kanokudom, Suvichada Assawakosri, Jira Chansaenroj, Donchida Srimuan, Thaksaporn Thatsanatorn, Siriporn Songtaisarana, Natthinee Sudhinaraset, Nasamon Wanlapakorn, Yong Poovorawan

## Abstract

**Background:** Severe acute respiratory syndrome-coronavirus-2 (SARS-CoV-2) infection can be asymptomatic in young children. Therefore, the true rate of infection is likely underestimated. Few data are available on the rate of infections in young children, and studies on the SARS-CoV-2 seroprevalence among children during omicron wave are limited. Our study aims to assess the SARS-CoV-2 infection-induced seroprevalence among children and estimated the associated risk factors for seropositivity.

**Methods:** A longitudinal serological survey was conducted from January 2021 through November 2022. Samples were tested for anti-nucleocapsid (N) IgG, anti-receptor binding domain (RBD) IgG using a chemiluminescent microparticle immunoassay (CMIA) and detected anti-RBD Immunoglobulin (Ig) using an electrochemiluminescence immunoassay (ECLIA). The vaccination and SARS-CoV-2 infection history were collected.

**Results:** A total of 452 serum samples were obtained from 249 children aged 5–7 years old who were annually followed-up in the longitudinal serological survey. Of these, 191 participants provided samples at two serial time points, including during the pre-and omicron dominant wave. Overall, seroprevalence induced by SARS-CoV-2 infection was increased from 9.1% (95%CI: 0.6-12.6%) during the pre-omicron wave to 49.7% (95%CI: 35.9-66.8%) during the omicron wave. Amongst seropositive individuals, the infection-induced seroprevalence was lower in vaccinated participants than those with no vaccination (40.4% vs. 57.4%; risk ratio, 0.71; 95%CI: 0.52–0.95). Nevertheless, the ratio of seropositive cases per recalled infection was 1.56 during the omicron dominant wave. In addition, overall seroprevalence induced by infection, vaccination and hybrid immunity was 76.6% (151/197; 95%CI: 54.6-97.9%) between January and November 2022.

**Conclusions:** our study reports an increase in infection-induced seroprevalence among children during the omicron wave. These findings highlight that estimating seroprevalence is crucial to monitor SARS-CoV-2 exposure, particularly in asymptomatic infection, and help to optimize public health policies and determine the effect of immunization in the pediatric population.

## Introduction

Most children with SARS-CoV-2 infection are asymptomatic at presentation. Thus, mildly symptomatic and asymptomatic children might not be tested for or diagnosed with SARS-CoV-2 infection [1]. As a result, pediatric COVID-19 cases are underestimated and likely higher than reported according to the reverse transcriptase-polymerase chain reaction (RT-PCR) results. Furthermore, children are commonly infected after close contact with other members in the same household. They are likely unaware of their infection status, potentially contributing to virus transmission [2].

Although children with COVID-19 typically develop a milder illness than adults, long-term complications including multisystem inflammatory syndrome in children (MIS-C) can occur after SARS-CoV-2 infection [3]. Recent evidence notes that COVID-19 vaccination in children is associated with a high level of protection against MIS-C and a reduced rate of hospitalization and deaths [4]. In Thailand, the Thai Food and Drugs Administration have approved BNT162b2 for vaccination in children aged 5–11 years old, and CoronaVac and BBIBP-CorV for children aged ≥6 years old. Consequently, data on seroprevalence in children is essential to inform and guide vaccine policies.

The value of a serological survey is the retrospective detection of individuals who had a previous infection, even in an asymptomatic case, which helps monitor virus transmission [5]. Anti-nucleocapsid (N) and anti-receptor binding domain (RBD) antibodies are elicited after natural infection or vaccination. However, anti-N antibodies can detect after receipt the inactivated vaccine but not detected after receipt of the virus vector or mRNA vaccines.

Moreover, some Thai children could elicit anti-N IgG because of being vaccinated with an inactivated vaccine. Consequently, the detection of anti-N and anti-RBD antibodies in unvaccinated individuals and anti-N antibodies in vaccinated individuals with mRNA vaccines could estimate the proportion of individuals who have been infected with SARS-CoV-2. In addition, serological surveys can inform the impact of immunization on the population.

Omicron was first detected in Thailand in mid-December 2021. The incidence of omicron infection rapidly increased and subsequently became the predominant variant by January 2022 [6]. Study demonstrated that the omicron variant is highly transmissible and can evade the immune system even in vaccinated or previously infected individuals [7]. In line with this, the spreading of omicron variants peaked between January and March 2022. In addition, infection by the omicron variant resulted in milder symptoms and was associated with lower risks of hospitalization and death than the delta variant [8]. However, few data are available on the rate of infections in young children, and studies on the SARS-CoV-2 seroprevalence among children during omicron wave are limited.

In this study, we assessed the seroprevalence of anti-SARS-CoV-2 antibodies induced by SARS-CoV-2 infection from January 2021 to November 2022 (the duration spanned the pre- and omicron wave) and estimated the associated risk factors for seropositivity. Data on seroprevalence can help to estimate the number of children who experience SARS-CoV-2 infection and assess the impact of immunization on protect SARS-CoV-2 infection in pediatric population which is paramount to establish public health prevention and vaccination strategies.

## Materials and methods

### Study design and sample collection

This study analyzed the serum samples from children who were followed-up in the longitudinal serological study of pertussis vaccine immunity at the Center of Excellence in Clinical Virology, Chulalongkorn Memorial Hospital, Bangkok, Thailand, from January 1, 2021 through November 9, 2022 [9]. Participants were invited to complete surveys on SARS-CoV-2 infection and COVID-19 vaccination. This survey included questions regarding previous SARS-CoV-2 infections detected by RT-PCR test or antigen test kit (ATK); date of infection; and vaccination data, including type, dose, and date of vaccination, as reported by the children’s parents. The survey responses were retrospectively collected between May and November 2022. The inclusion criteria were the same as the longitudinal serological study of the pertussis vaccine immunity. However, the exclusion criteria were parents who did not want to disclose vaccination and infection history, and children who previously received any dose of the inactivated COVID-19 vaccine, because such individuals could elicit anti-N IgG from the inactivated vaccine.

The Research Ethics Committee of the Faculty of Medicine, Chulalongkorn University approved the study (IRB number: 173/63). The study protocol adhered to the tenets of the Declaration of Helsinki and Good Clinical Practice principles. Written informed consent was obtained from the parents or legal guardians of all participating children.

### Serological analysis

Sera samples collected from eligible participants were subjected to IgG-specific testing against SARS-CoV-2 nucleocapsid (N) protein using a chemiluminescent microparticle immunoassay (CMIA) (Abbott Architect Immunoassay; Abbott Diagnostics, Abbott Park, IL, USA). According to the manufacturer’s instructions, we considered samples with a signal-to-cut-off ratio ≥1.4 as anti-N IgG positive, while samples with a value <1.4 was considered as seronegative. The total immunoglobulin (Ig) and IgG specific against SARS-CoV-2 receptor binding domain (RBD) quantitatively measured using the Roche Elecsys anti-SARS-CoV-2 immunoglobulin immune assay (Roche) and SARS-CoV-2 Quant IgG II (Abbott Diagnostics, Abbott Park, IL, USA), respectively. Antibody level is expressed as U/mL for anti-RBD Ig, with a cut-off ≥ 0.8 defined as positive and BAU/mL for anti-RBD IgG, with values ≥7.1 scored positive.

### Classify the samples with seropositivity

Infection-induced seropositivity was estimated based on the presence of anti-RBD Ig (≥ 0.8 U/mL), anti-RBD IgG (≥7.1 BAU/mL) or anti-N IgG (≥ 1.4 S/C) among unvaccinated children and the presence of anti-N IgG or having an evidence of SARS-CoV-2 infection during omicron variants in vaccinated individuals with BNT162b2 vaccine. Vaccination status was classify according to the dates of reported infection and vaccination. Overall seroprevalence induced by infection, vaccination, and hybrid immunity was estimated based on the anti-RBD antibody seropositivity in both unvaccinated and vaccinated participants.

### Statistical analysis

Data are mainly presented as numbers and percentages. The primary outcome was infection-induced seroprevalence. Risk ratios associated with infection-induced seropositivity and demographic characteristics were calculated using the chi-square test. The ratio of children with infection-induced seropositivity during the omicron dominant wave per recalled of previous SARS-CoV-2 infection were assessed. Statistical analysis was conducted using SPSS v23.0 (IBM Corp, Armonk, NY, USA). A *p*-value <0.05 was considered to indicate statistically significant differences.

## Results

### Study participants

There were 452 samples from 249 participants enrolled in the source study (Fig 1). Of these, 241 samples were collected from January through December 2021 (hereafter referred to as the pre-omicron wave), and 211 samples were collected from January to November 2022 (hereafter referred to as the omicron-dominant wave). Between January and November 2022, two participants were excluded because they lacked vaccination history, and 12 participants were excluded because they received an inactivated vaccine. Thus, 438 serum samples from 247 participants were eligible and met the criteria for the final analysis.

**Figure 1.**
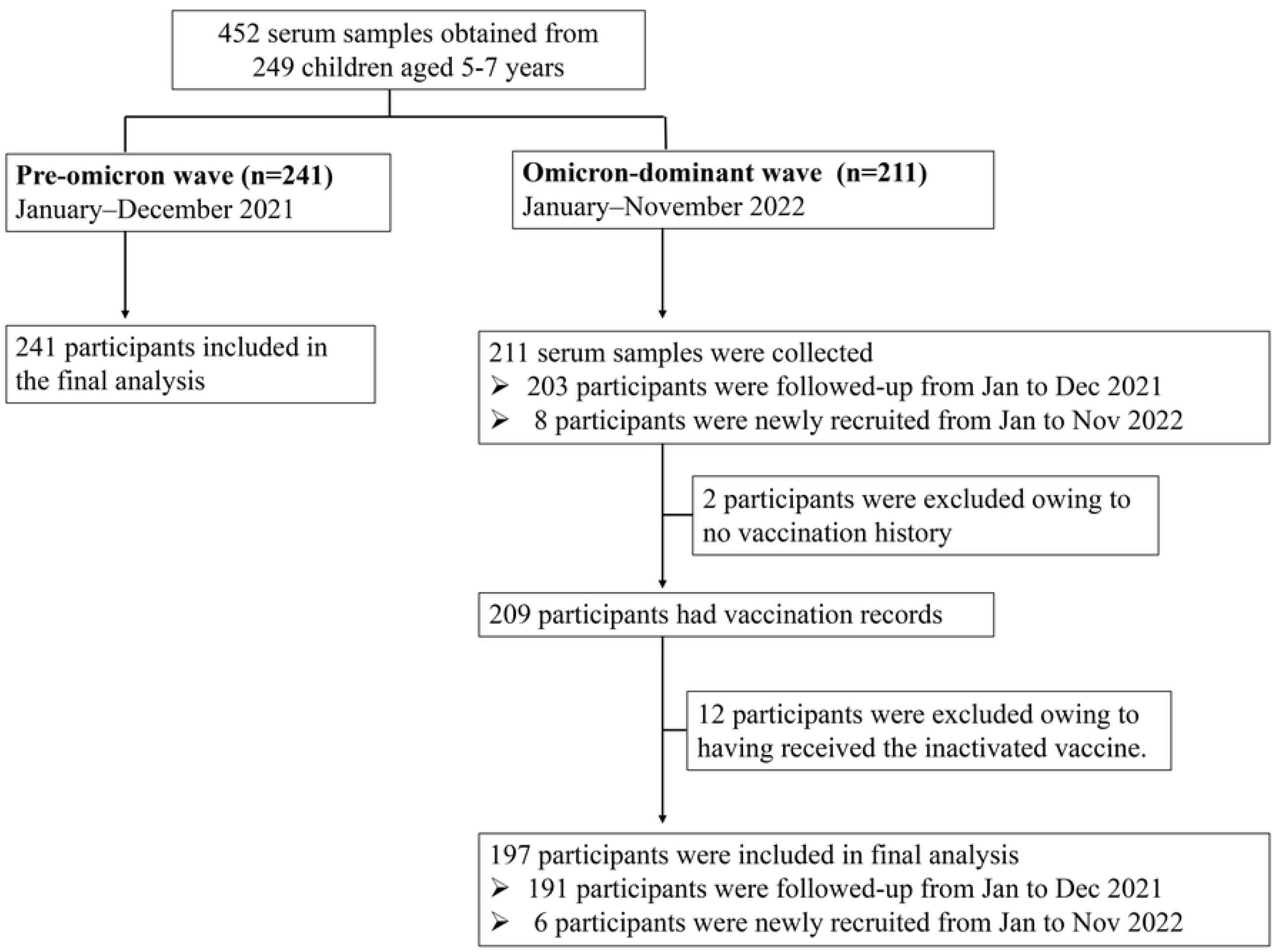
Recruitment of participants.

In total, 438 samples were obtained, including 55% (241/438) serum samples collected between January and December 2021 and 45% (197/438) collected between January and November 2022, during which time omicron was predominantly circulating. The mean (SD) age of study participants enrolled during pre- and omicron wave was 5.12 (0.34) and 6.16 (0.36) years old, and there were 51% female and 49% male subjects (S1 Table). Of these, 191 participants provided samples at two serial time points, including during the pre-and omicron dominant wave, and the median interval between the two blood samplings was 362 days (IQR: 343–376). During the omicron-dominant wave, 45.2% (89/197) of participants received at least one dose of BNT162b2 by February 2022 included 42.7% (38/89), 55.1% (49/89) and 2.2% (2/89) of participants who had been received a single dose, two-doses and three doses of BNT162b2, respectively, while no participant enrolled during the pre-omicron wave received the COVID-19 vaccine.

### Seroprevalence induced by SARS-CoV-2 infection

There were 120 samples out of a total of 438 that tested anti-RBD Ig or anti-N IgG seropositive for unvaccinated participants and anti-N IgG seropositive or having reported of SARS-CoV-2 infection during omicron for vaccinated participants, resulting in an estimated infection-induced seroprevalence of 27.4% (95%CI: 15.7–40.3%) (Fig 2). The infection-induced seroprevalence increased from 9.1% (95%CI: 0.6–12.6%) for January to December 2021 to 49.7% (95%CI: 35.9–66.8%) observed between January and November 2022. Amongst seropositive participants during the pre-omicron wave, 14 participants were follow-up the serological cohorts during the omicron-dominant wave and 87.5 % (14/16) of children were provided evidence of re-infection.

**Figure 2.**
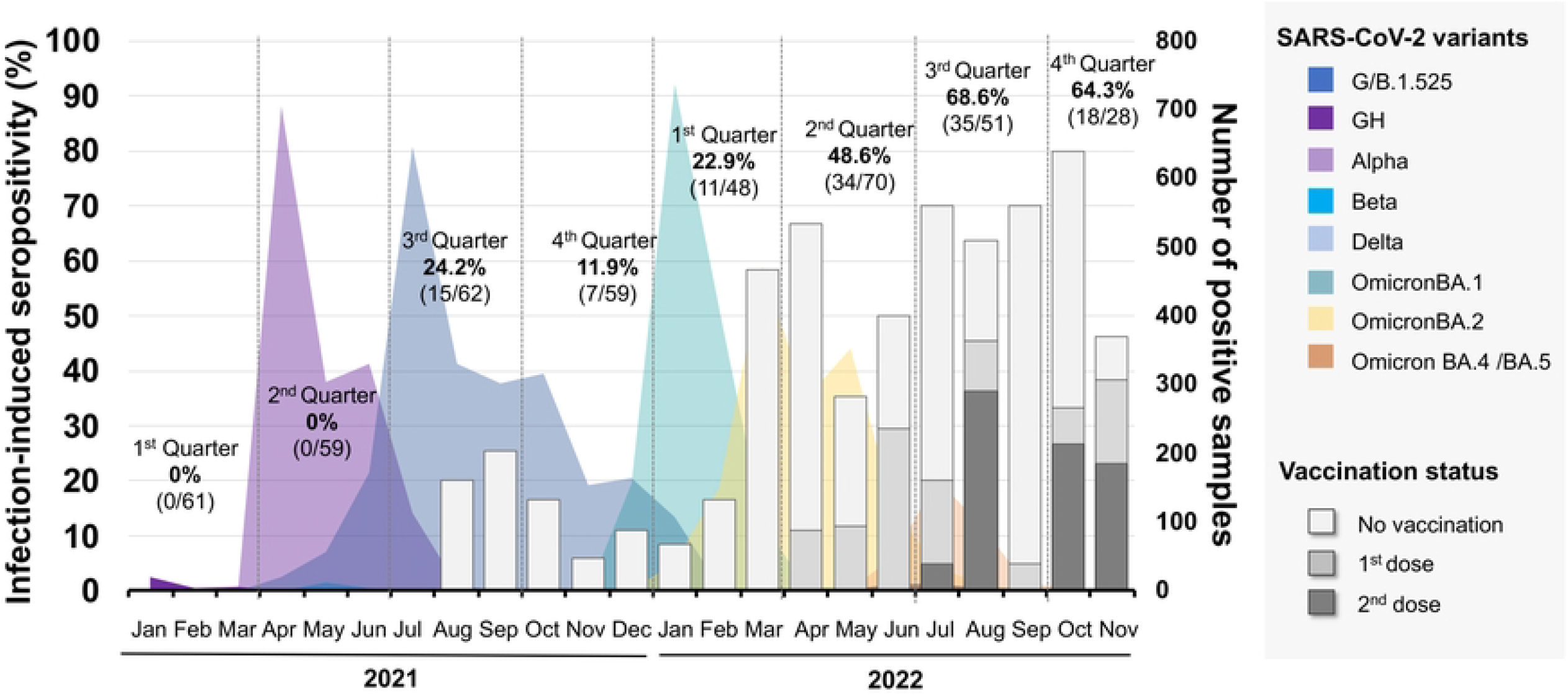
Monthly distribution of SARS-CoV-2 variants and infection-induced seroprevalence. SARS-CoV-2 infection-induced seropositivity was identified by the seropositivity of anti-N IgG or anti-RBD antibody for unvaccinated individuals and anti-N IgG seropositivity or having reported of SARS-CoV-2 infection for vaccinated individuals with BNT162b2 vaccine. Sera samples were collected from Thai children between January 2021 and November 2022. Data on SARS-CoV-2 variants circulating during the study period was obtained from a previous report [6]. The stacked bar graph represents the percentage of infection-induced seropositivity according to vaccination status (see left Y-axis). Color shaded areas indicate the number of positive samples of SARS-CoV-2 variants (see right Y -axis). The vertical dotted lines indicate the quarter.

### Factors associated with infection-induced seropositivity

Among unvaccinated participants, samples collected during the omicron dominant wave were more likely seropositive than those collected during the pre-omicron wave (57.4% vs. 9.1%; risk ratio, 6.29; 95%CI: 4.09–9.67, p<0.001) (Table 1). Notably, vaccinated participants was less likely infected with SARS-CoV-2 than unvaccinated participants (40.4% vs. 57.4%; risk ratio, 0.71; 95%CI: 0.52–0.95; p<0.05) and participants vaccinated with two-doses of BNT162b2 were associated with a low seroprevalence than those immunized with a single dose (24.5% vs. %; risk ratio, 0.39; 95%CI: 0.22–0.67, p<0.001). There was no difference in seroprevalence rate between participants living in a household with ≥5 members and those living in a household with <5 members during the omicron wave. However, children living with household members who had previous COVID-19 infection had a higher risk of seropositivity than those living with household members without SARS-CoV-2 infection (73.7% vs.21.1%; risk ratio, 3.45; 95%CI: 2.19–5.55; P<0.001).

**Table 1.** SARS-CoV-2 infection-induced seropositivity and risk factors for seropositivity.

| Characteristic | Overall | Infection-induced seroprevalence |  | Risk ratio associated with seropositivity, |
| --- | --- | --- | --- | --- |
|  | n | n | (%) | Risk ratio (95%CI) |
| Overall | 438 | 120 | 27.4% | - |
| -Pre-omicron wave | 241 | 22 | 9.1% | Ref |
| -Omicron dominant wave | 197 | 98 | 49.7% | 5.45 (3.57–8.31)*** |
| Sex, n (%) according to the study period |  |  |  |  |
| Pre-omicron wave |  |  |  |  |
| Boy | 117 | 12 | 10.3% | Ref |
| Girl | 124 | 10 | 8.1% | 0.79 (0.35–1.75) |
| Omicron-dominant wave |  |  |  |  |
| Boy | 96 | 49 | 51.0% | Ref |
| Girl | 101 | 49 | 48.5% | 0.95 (0.72–1.26) |
| Vaccination status according to<br>dates of infection and<br>vaccination |  |  |  |  |
| Unvaccinated |  |  |  |  |
| Pre-omicron wave | 241 | 22 | 9.1% | Ref |
| Omicron-dominant wave | 108 | 62 | 57.4% | 6.29 (4.09–9.67) *** |
| Omicron dominant wave<br>(n=197) |  |  |  |  |
| Unvaccinated | 108 | 62 | 57.4% | Ref* |
| Vaccination | 89 | 36 | 40.4% | 0.71 (0.52–0.95) |
| Dose of vaccine |  |  |  |  |
| -BNT162b2 (1 dose) | 38 | 24 | 63.2% | Ref |
| -BNT162b2 (2 doses) | 49 | 12 | 24.5% | 0.39 (0.22–0.67)*** |
| -BNT162b2 (3 doses) | 2 | 0 | 0% | - |
| Pre-omicron wave |  |  |  |  |
| Number of household members,<br>n |  |  |  |  |
| 2–4 | 96 | 4 | 4.2% | Ref |
| ≥5 | 62 | 12 | 19.4% | 4.65 (1.57-13.76)** |
| N/A | 83 | 6 | 7.2 % | - |
| Omicron-dominant wave |  |  |  |  |
| Number of household members, n |  |  |  |  |
| 2–4 | 100 | 50 | 50.0% | Ref |
| ≥5 | 64 | 35 | 54.7% | 1.01 (0.81–1.47) |
| N/A | 33 | 13 | 39.4% | - |
| Confirmed COVID-19 infection in household members tested via PCR or ATK during the omicron wave, n |  |  |  |  |
| No | 71 | 15 | 21.1% | Ref |
| Yes | 95 | 70 | 73.7% | 3.45 (2.19–5.55)*** |
| N/A | 31 | 13 | 41.9% | - |

Statistical analysis was performed using the chi-square test. \* indicates $p < 0.05$ , \*\*\*indicates $p < 0.001$ \*\*\*indicates $p < 0.001$ .

#### Infection-induced seropositivity associated with previous SARS-CoV-2 testing

We further estimated the ratio of infection-induced seropositive cases per recalled infection during the omicron dominant wave. Overall, 89.4% (176/197) participants completed a survey on infection history. Subsequently, 92 of 176 children showed infection-induced seropositivity. Of these, 64.1% (59/92) previously reported SARS-CoV-2 infection, and the median interval between dates of infection and blood collection was 95 days (IQR: 64–175), while 35.9% (33/92) reported being unaware of their infection status (Table 2). Therefore, the ratio of infection-induced seropositive cases per recalled infection during omicron was 1.56 (92/59). Moreover, only 4.3% (4/94) seronegative participants reported previous SARS-CoV-2 infection, for which the median interval between infection and blood collection dates was 311 days (IQR: 272.5–316.5).

**Table 2.** Association between infection-induced seropositivity and previous SARS-CoV-2 infection. Number of children who complete a survey of infection history (n=176) and infection-induced seropositivity during the omicron-dominant wave (January–November 2022).

|  | Infection-induced<br>seropositive (n=92) | Infection-induced<br>seronegative (n=94) |
| --- | --- | --- |
| History of previous infection<br>(n=63) | 59 | 4 |
| Interval between the reported date<br>of infection and blood collection,<br>median (IQR) | 95 (64–175) | 311 (272.5–316.5) |
| No history of previous infection<br>(n=113) | 33 | 80 |

Previous SARS-CoV-2 infection was diagnosed by RT-PCR or ATK test

### Seroprevalence induced by infection, vaccination and hybrid immunity

We further estimated the overall seroprevalence induced by natural infection, vaccination and hybrid immunity based on the seropositivity of anti-RBD antibodies (Fig 3). Regarding the vaccine implementation in children, the seropositivity of anti-RBD antibodies was 76.6% (151/197; 95%CI: 54.6-97.9%) between January and November 2022 which increased by Mar 2022 and reached greater than 80% (95%CI: 88.6-96.2%) since April 2022.

**Figure 3.**
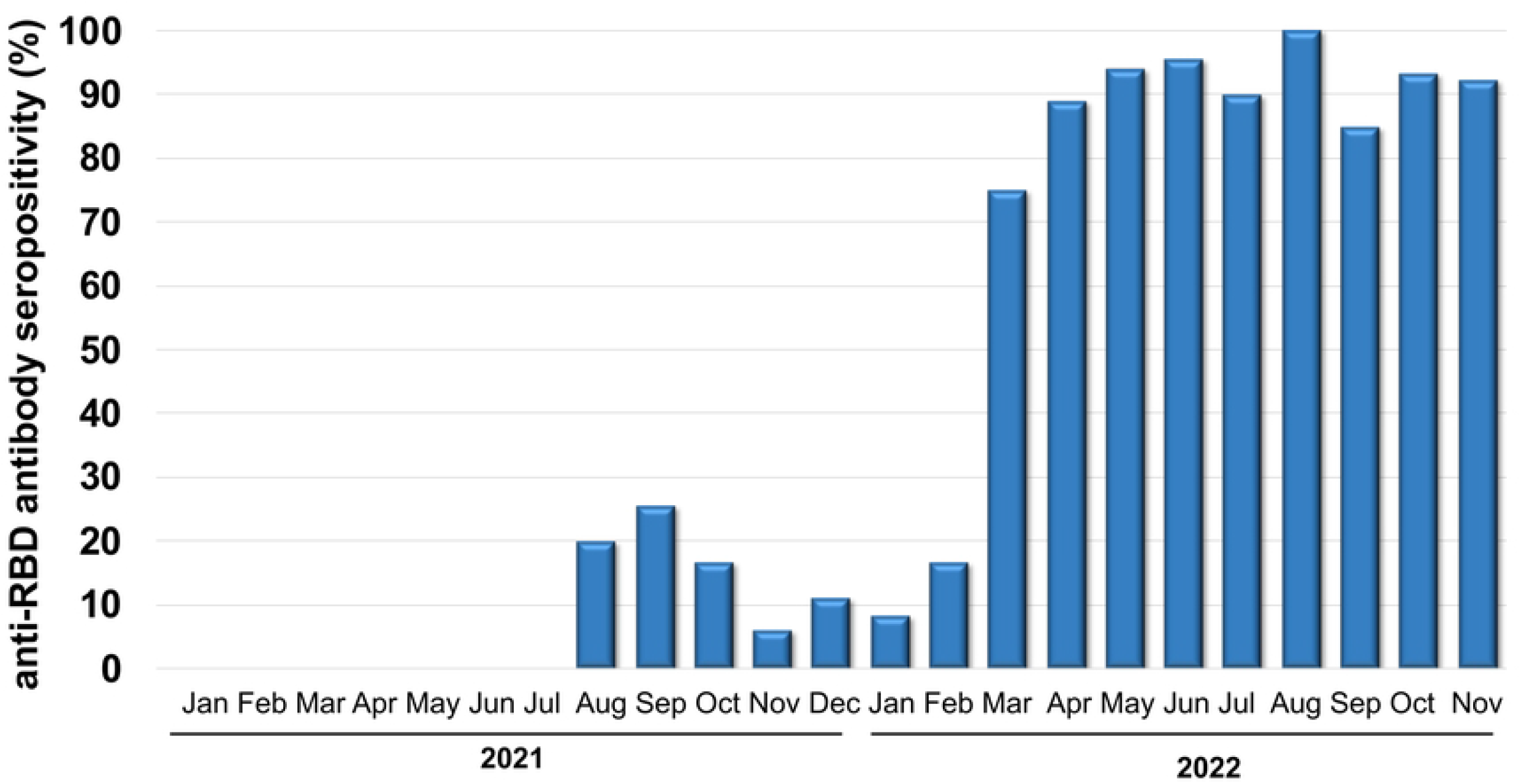
Monthly distribution of seroprevalence induced by infection, vaccination, and hybrid immunity observed in children aged 5-7 years old between January 2021 and November 2022. Seroprevalence induced by infection, vaccination, and hybrid immunity was estimated based on the anti-RBD antibody seropositivity in both unvaccinated and vaccinated participants.

## Discussion

Our findings report the increase in seroprevalence induced by infection among children from January through November 2022, indicating the high rates of SARS-CoV-2 infection during the omicron wave. This is in agreement with other studies that have reported a significant rise in the seroprevalence rate among children during the omicron wave in South Africa [10] and the United States [11]. This finding supported the previous report which showed that the omicron variant is more transmissible than the previous SARS-CoV-2 variants [7].

Our results also showed that even children who received the BNT162b2 vaccine were anti-N IgG seropositive during the omicron wave. This result might be related to the antibody evasion by omicron variants and the limit of vaccine efficacy among children [12]. Although vaccinated children were susceptible to being infected by the omicron variant, the risks of infection and hospitalization were reduced for vaccinated compared to unvaccinated children aged between 5 and 11 years [13]. In addition, our study showed that vaccinated children with two-dose vaccines were less likely to be seropositive than those who were vaccinated with a single dose. This result was supported by a meta-analysis study that suggested that the estimated mean secondary attack rates observed during the omicron wave were higher in partially vaccinated cases (76.8%, 95%CI: 7.7%–99.2%) than in fully vaccinated cases (50.8%, 95%CI: 47.9%–53.8%) [14].

Although an increased number of household members did not affect the risk of seropositivity in children, we found a higher risk of seropositivity was observed in children living with infected household members than those living with non-infected household members. This finding was consistent with a study from Switzerland that indicated the risk of being seropositive in children aged <6 years was more likely associated with the number of household members who tested positive for SARS-CoV-2, indicating the SARS-CoV-2 transmission for children aged <6 years occurs commonly within the family [15].

The proportion of anti-N IgG or anti-RBD antibody seropositive among unvaccinated and anti-N IgG seropositive among vaccinated cases may reflect the true rate of infections, including mildly symptomatic or asymptomatic cases and undetected by COVID-19 testing. Our study found that 35.9% children aged 5–7 years with seropositivity reported no previous infection.

This result may reflect either asymptomatic infection or undetected infection upon COVID-19 testing in patients likely unaware of their infection status, in which case the virus remains actively transmissible [16]. In addition, our estimated ratio of seropositive cases per recall infection was lower than that reported in a seroprevalence study in children aged 3–11 years from Germany between March and May 2021 (1.56 vs. 3.0) [17] and another study of awareness in adults during the omicron wave (1.56 vs. 2.28) [5].

The anti-N IgG seronegative results in children that reported infection may be owing to the long interval from infection to blood sampling and the severity of disease after infection. This result was supported by a previous study in adults indicating that 87.5% individuals had detectable the anti-N IgG even 3 months post-infection, while only 26.6% individuals had detectable the anti-N IgG 12 months post-infection [18]. In addition, recent evidence showed that the sensitivity of anti-N IgG detection among individuals with recent infection defined by RT-PCR testing ranged from 74% to 81% during the omicron wave [19]. To overcome the limitation of anti-N IgG detection, our study included anti-RBD antibodies to classify the seropositivity induced by SARS-CoV-2 infection in unvaccinated participants and be considered evidence of SARS-CoV-2 infection in vaccinated participants.

The limitations of the study include the small sample size, as this may not represent the seroprevalence in the general population. Further, the present study could have underestimated the cumulative seropositivity because children with inactivated vaccines were not included. Furthermore, anti-N antibody titers waned over time, and the sensitivity of the anti-N IgG assay to detect the antibody was dependent on the time between the infection and blood collection and vaccination status [20].

In conclusion, our study reports an increase in infection-induced seroprevalence among children during the omicron wave and overall seroprevalence induced by natural infection, vaccination and hybrid immunity. This shows that the benefit of anti-SARS-CoV-2 antibody detection can help to assess the seroprevalence induced by infection which subsequently track the transmission of SARS-CoV-2 variants and determine the effect of immunization in the pediatric population.

## Data Availability

All relevant data are within the manuscript and its Supporting Information files.

## Acknowledgments

We would like to thank all the staff of the Center of Excellence in Clinical Virology. This research was financially supported by the Health Systems Research Institute (HSRI), National Research Council of Thailand (NRCT), the Center of Excellence in Clinical Virology, Chulalongkorn University, and King Chulalongkorn Memorial Hospital, MK Restaurant Group Aunt Thongkam Foundation, BJC Big C Foundation, and the Second Century Fund (C2F), Chulalongkorn University.

## Author Contributions

Conceptualization, N.S.(Nungruthai Suntronwong), N.W. and Y.P.; data curation, N.S. (Nungruthai Suntronwong), J.P., D.S., T.T., S.S., and N.S. (Natthinee Sudhinaraset); formal analysis, N.S.(Nungruthai Suntronwong); methodology, P.V., S.K. (Sirapa Klinfueng), S.K. (Sitthichai Kanokudom), S.A., and J.C.; project administration, Y.P.; writing— original draft, N.S. (Nungruthai Suntronwong); writing—review and editing, N.S. (Nungruthai Suntronwong), S.K. (Sitthichai Kanokudom), N.W., and Y.P. All authors have read and agreed to the published version of the manuscript.

## Institutional Review Board Statement

The study protocol was approved by the Research Ethics Committee of the Faculty of Medicine, Chulalongkorn University (IRB number: 173/63). The study protocol adhered to the tenets of the Declaration of Helsinki and Good Clinical Practice principles. Written informed consent was obtained from the parents or legal guardians of all participating children.

## Data Availability Statement

All relevant data are within the manuscript and its Supporting Information files.

## Supplementary Information

**S1 Table. Baseline characteristics and data summaries of vaccination status and infection history**. The final study participants recruited from January to December 2021 (pre-omicron wave) and January to November 2022 (omicron-dominant wave). Sera samples were categized according to the dates of blood collection into pre- and omicron dominant wave. Vaccination status was classify according to the dates of reported infection and vaccination.

